# Temporal Variations in the Intensity of Care Provided to Community and Nursing Home Residents Who Died of COVID-19 in Ontario, Canada

**DOI:** 10.1101/2020.11.06.20227140

**Authors:** Kevin A. Brown, Nick Daneman, Sarah A. Buchan, Adrienne K. Chan, Nathan M. Stall

**Affiliations:** Public Health Ontario, Toronto, Canada; Department of Medicine, University of Toronto, Canada

## Abstract

Introduction - Worldwide, nursing home residents have experienced disproportionately high COVID-19 mortality due to the intersection of congregate living, multimorbidity, and advanced age. Among 12 OECD countries, Canada has had the highest proportion of COVID-19 deaths in nursing home residents (78%), raising concerns about a skewed pandemic response that averted much transmission and mortality in community-dwelling residents, but did not adequately protect those in nursing homes. To investigate this, we measured temporal variations in hospitalizations among community and nursing home-dwelling decedents with COVID-19 during the first and second waves of the pandemic.

Methods - We conducted a population-based cohort study of residents of Ontario, Canada with COVID-19 who died between March 11, 2020 (first COVID-19 death in Ontario) and October 28, 2020. We examined hospitalization prior to death as a function of 4 factors: community (defined as all non-nursing home residents) vs. nursing home residence, age in years (<70, 70-79, 80-89, ≥90), gender, and month of death (1st wave: March-April [peak], May, June-July 2020 [nadir], 2nd wave: August-October 2020).

Results - A total of 3,114 people with confirmed COVID-19 died in Ontario from March to October, 2020 (Table 1), of whom 1,354 (43.5%) were hospitalized prior to death (median: 9 days before death, interquartile range: 4-19). Among nursing home decedents (N=2000), 22.4% were admitted to hospital prior to death, but this varied substantially from a low of 15.5% in March-April (peak of wave 1) to a high of 41.2% in June-July (nadir of wave 1). Among community-dwelling decedents (N=1,114), admission to acute care was higher (81.4%) and remained relatively stable throughout the first and second waves. Similar temporal trends for nursing home versus community decedents were apparent in age-stratified analyses (Figure 1). Women who died were less likely to have been hospitalized compared to men in both community (80% women vs 84% men) and nursing home (21% women vs 24% men) settings.

Discussion - Only a minority of Ontario nursing home residents who died of COVID-19 were hospitalized prior to death, and that there were substantial temporal variations, with hospitalizations reaching their lowest point when overall COVID-19 incidence peaked in mid-April, 2020. While many nursing home residents had pre-pandemic advance directives precluding hospitalization, the low admission rate observed in March-April 2020 (15.5%) was inconsistent with both higher admission rates in subsequent months (>30%), and comparatively stable rates among community-dwelling adults. Our findings substantiate reports suggesting that hospitalizations for nursing home residents with COVID-19 were low during the peak of the pandemic’s first wave in Canada, which may have contributed to the particularly high concentration of COVID-19 mortality in Ontario’s nursing homes.

## Introduction

Worldwide, nursing home residents have experienced disproportionately high COVID-19 mortality due to the intersection of congregate living, multimorbidity, and advanced age.^1^ Among 12 OECD countries, Canada has had the highest proportion of COVID-19 deaths in nursing home residents (78%), raising concerns about a skewed pandemic response that averted much transmission and mortality in community-dwelling residents, but did not adequately protect those in nursing homes.^2^ In Canada’s most populous province of Ontario there was no official policy denying hospitalizations for nursing home residents with COVID-19, yet media reports and a provincial commission suggest that resident transfers to hospital were strongly discouraged at the onset of the pandemic.^3,4^ To investigate this, we measured temporal variations in hospitalizations among community and nursing home-dwelling decedents with COVID-19 during the first and second waves of the pandemic.

## Methods

We conducted a population-based cohort study of residents of Ontario, Canada with COVID-19 who died between March 11, 2020 (first COVID-19 death in Ontario) and October 28, 2020. Data were extracted on the last day of the study period from the provincial reportable disease surveillance system, which records all reported COVID-19 cases, and their outcomes. All Ontario residents receive medically necessary services under a publicly funded provincial health insurance program, and nursing home residents receive additional personal and nursing care. We examined hospitalization prior to death as a function of 4 factors: community (defined as all non-nursing home residents) vs. nursing home residence, age in years (<70, 70-79, 80-89, ≥ 90), gender, and month of death (1^st^ wave: March-April [peak], May, June-July 2020 [nadir], 2^nd^ wave: August-October 2020). The study received a waiver of consent from the research ethics board of Public Health Ontario.

## Results

A total of 3,114 people with confirmed COVID-19 died in Ontario from March to October, 2020 (Table), of whom 1,354 (43.5%) were hospitalized prior to death (median: 9 days before death, interquartile range: 4-19). Among nursing home decedents (N=2000), 22.4% were admitted to hospital prior to death, but this varied substantially from a low of 15.5% in March-April (peak of wave 1) to a high of 41.2% in June-July (nadir of wave 1). Among community-dwelling decedents (N=1,114), admission to acute care was higher (81.4%) and remained relatively stable throughout the first and second waves. Similar temporal trends for nursing home versus community decedents were apparent in age-stratified analyses (Figure). Women who died were less likely to have been hospitalized compared to men in both community (80% women vs 84% men) and nursing home (21% women vs 24% men) settings.

**Table.** Admissions to hospital prior to death among community and nursing home residents who died with COVID-19, March to October 2020 (N=3,114).

|  | Community Residents |  |  | Nursing Home Residents |  |  |
| --- | --- | --- | --- | --- | --- | --- |
|  | N | Hospitalized<br>prior to death<br>(N [%]) | p-value* | N | Hospitalized<br>prior to death<br>(N [%]) | p-value* |
| Total | 1,114 | 907 (81.4) |  | 2,000 | 447 (22.4) |  |
| Month |  |  |  |  |  |  |
| March-April | 532 | 426 (80.1) |  | 1,028 | 159 (15.5) |  |
| May | 306 | 253 (82.7) | <0.001 | 729 | 196 (26.9) | <0.001 |
| June-July | 143 | 127 (88.8) |  | 165 | 68 (41.2) |  |
| August-October | 133 | 101 (75.9) |  | 78 | 24 (30.8) |  |
| Age (years) |  |  |  |  |  |  |
| <70 | 287 | 239 (83.3) |  | 141 | 56 (39.7) |  |
| 70-79 | 250 | 217 (86.8) | <0.001 | 299 | 82 (27.4) | <0.001 |
| 80-89 | 352 | 291 (82.7) |  | 778 | 175 (22.5) |  |
| ≥90 | 225 | 160 (71.1) |  | 782 | 134 (17.1) |  |
| Gender* |  |  |  |  |  |  |
| Male | 621 | 517 (83.3) | 0.1 | 811 | 202 (24.9) | 0.03 |
| Female | 491 | 390 (79.4) |  | 1,163 | 242 (20.8) |  |

\* P-values for differences in proportions were based on Pearson's chi-squared test
\*\* Deaths with unknown gender are not presented below due to small cell counts (N=28)

**Figure.**
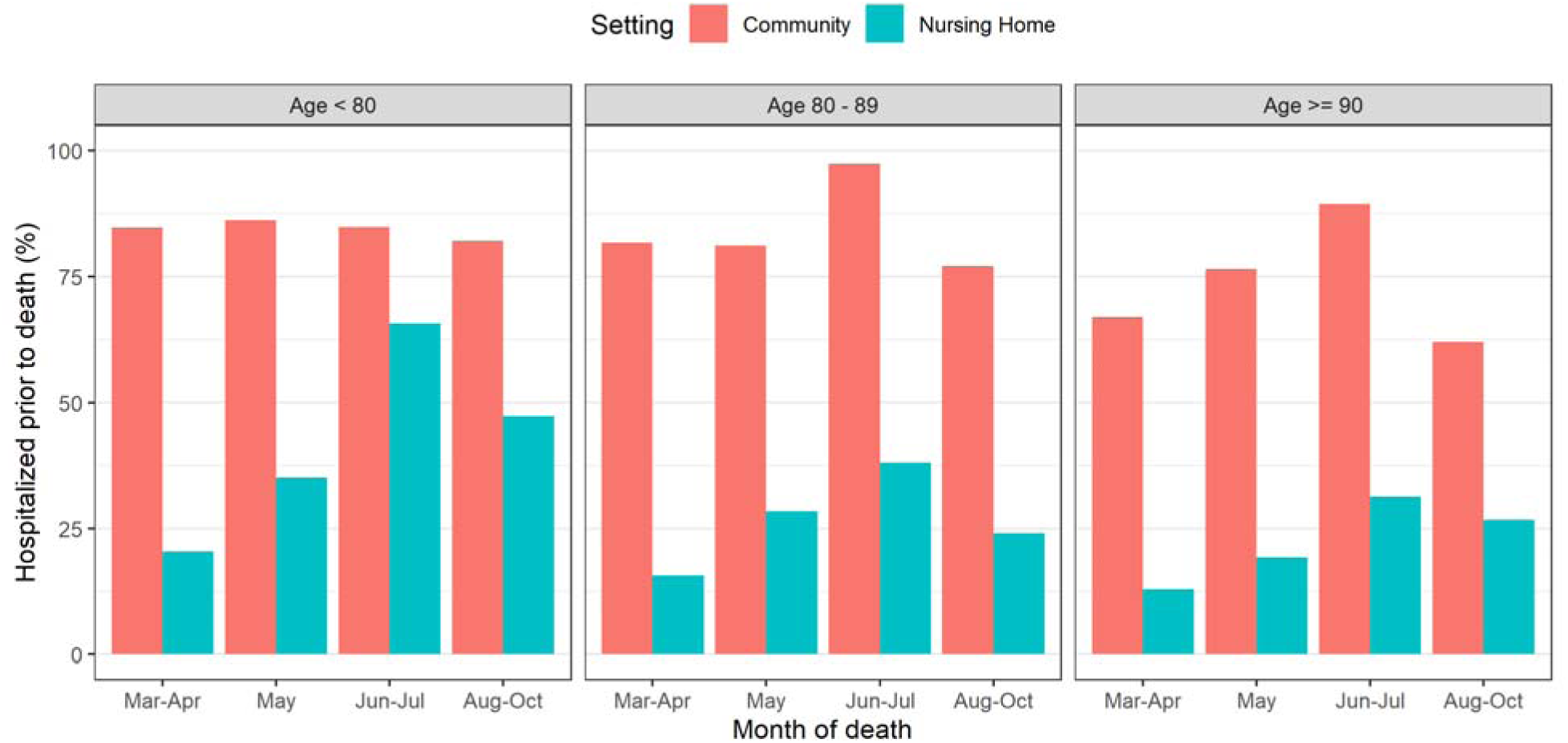
Admissions to hospital prior to death among community and nursing home decedents with COVID-19, March to October 2020, (N=3,114), stratified by age group (<80, 80-89, ≥90).

## Discussion

Only a minority of Ontario nursing home residents who died of COVID-19 were hospitalized prior to death, and that there were substantial temporal variations, with hospitalizations reaching their lowest point when overall COVID-19 incidence peaked in mid-April, 2020. While many nursing home residents had pre-pandemic advance directives precluding hospitalization, the low admission rate observed in March-April 2020 (15.5%) was inconsistent with both higher admission rates in subsequent months (>30%), and comparatively stable rates among community-dwelling adults. This raises concerns for other jursidictions, including the United States, which has seen substantial inter-state variations in the proportion of COVID-19 deaths occurring in nursing homes, ranging from 18% in Nevada to 79% in New Hampshire.^5^

This analysis has limitations. Hospitalizations recorded in the surveillance system may undercount hospitalizations compared to administrative records (sensitivity=0.87, specificity=0.98).^6^ We also lacked information on advance directives which may have impacted the decision to hospitalize.

Our findings substantiate reports suggesting that hospitalizations for nursing home residents with COVID-19 were low during the peak of the pandemic’s first wave in Canada, which may have contributed to the particuarly high concentration of COVID-19 mortality in Ontario’s nursing homes.

## Data Availability

The specific dataset used for this analysis is not publically available. Ontario's Ministry of Health does make a modified version of this data publicly available (https://covid-19.ontario.ca/data). Questions regarding access to data should be directed to the ministry of health through the above website.

## References

1. Ouslander JG, Grabowski DC. COVID□19 in Nursing Homes: Calming the Perfect Storm. J Am Geriatr Soc. 2020;68(10):2153–2162. doi:10.1111/jgs.16784

2. Sepulveda ER, Stall NM, Sinha SK. A Comparison of COVID-19 Mortality Rates among Long-Term Care Residents in 12 OECD Countries. Journal of the American Medical Directors Association. Published online September 2020:S152586102030791X. doi:10.1016/j.jamda.2020.08.039

3. Reith T. As COVID-19 rips through nursing homes, some tell loved ones there’s “no benefit” to sending ill to hospital | CBC News. CBC. Published April 3, 2020. Accessed October 28, 2020. https://www.cbc.ca/news/health/covid-19-long-term-care-1.5519657

4. Ontario Long Term Care Commission. Meeting with Chartwell Retirement Residences.; 2020:93. http://www.ltccommission-commissionsld.ca/transcripts/pdf/Chartwell_Retirement_Residences_Transcript_October_09_2020.pdf

5. About 38% of U.S. Coronavirus Deaths Are Linked to Nursing Homes. The New York Times. https://www.nytimes.com/interactive/2020/us/coronavirus-nursing-homes.html. xPublished June 27, 2020. Accessed October 29, 2020.

6. Chung H. Validation of COVID-19 Outcomes in Public Health Surveillance Data using Health Administrative Databases. Personal e-mail communication. October 21, 2020.

